# Role of the HIV-sensitive protection services in mitigating the challenges and vulnerability of the children affected by HIV/AIDS in Bangladesh: a qualitative study

**DOI:** 10.1101/2020.07.21.20158824

**Authors:** Tahmina Afroz, Suborna Camellia, Tajudeen Oyewale, M. Ziya Uddin, Ilias Mahmud

## Abstract

Children are particularly vulnerable to the consequences of HIV/AIDS. HIV-sensitive protection services (HSPS) can reduce vulnerability by increasing access to HIV/AIDS prevention, treatment, and basic social protection services. Few organizations in Bangladesh offer these services to children affected by HIV/AIDS (CABA). This paper reports on the challenges faced by CABA due to their or their parents’ HIV/AIDS status and the role of HSPS on mitigating these challenges. We did fifteen in-depth interviews with purposively chosen CABA, and seventeen in-depth interviews with purposively chosen caregivers of CABA. We found, CABA faced significant challenges such as poverty, emotional stress, stigma in their lives. Gender norms increased these vulnerabilities especially among women. HSPS services helped through motivating parents or guardians to send their children to school and thus reduce child marriage and child labour. Also, helped in improving children’s self-esteem with improving relationships between children and parents. Though the services were available, accessibility was limited due to distance, roundtrip cost, poor health conditions of the parents and gender issues.HSPS are effective in improving children’s well-being. Continued and increased support to access services including financial assistance, psychosocial counselling, and community sensitization activities is recommended.

## Introduction

Children are particularly vulnerable to HIV (Human Immunodeficiency Virus)/AIDS (Acquired Immune Deficiency Syndrome) in Bangladesh (Islam et al., 2014). Among the infected population, 13% are adolescents aged below 20 years (NASP et al., 2016). However, only 12% of young women (NIPORT et al., 2016) and 14.4% of young men (NIPORT et al., 2013) have comprehensive knowledge of AIDS prevention in Bangladesh. Utilization of AIDS prevention, treatment and care services is also low (UNICEF, 2009, 2014). Only 28% of HIV positive pregnant mothers are receiving antiretroviral (ARV) for prevention of mother to child transmission (PMTCT) (UNAIDS, 2018).

Children affected by HIV/AIDS (CABA) are children under the age of 18 years who are HIV infected or whose parent/s or caregiver/s are HIV positive or have died due to AIDS (SAARC & UNICEF, 2008). There are 2,389 CABA in Bangladesh and 54% of all documented CABA were aged 10 – 18 years (NASP et al., 2013). According to a 2016 report, among the new cases of HIV, 5.5% were children (NASP et al., 2016).

After losing a parent, AIDS orphans often struggle to gain access to necessities and social services. They often must support themselves financially, which in some cases forces them into child labour (UNICEF, 2006). Even before they are orphans, the children of people living with HIV/AIDS (PLHIV) often experience family instability and neglect (Serey et al., 2011; Curley et al., 2010; Walker et al., 2011).

The stigma, shame, isolation, and fear that CABA feel can have a disastrous effect on their psychosocial wellbeing. To address these challenges, CABA need a wide variety of social protection services. Social protection services help in improving health and accessing education by reducing stigma and discrimination (UNICEF, 2017). Few organizations in Bangladesh were providing HIV-sensitive protection services (HSPS) to access to health, education and nutrition services for CABA (Azim et al, 2008). However, it is crucial to understand the role of HSPS on addressing the challenges faced by CABA. This paper reports on the challenges that were faced by CABA due to their or their parents’ HIV/AIDS status and the role of HSPS on mitigating these challenges.

## Materials and methods

### Study design

This study adopted a qualitative design to understand the challenges and vulnerability of the children affected by HIV/AIDS in Bangladesh and the role of the HIV-sensitive protection services in mitigating these.

### Study settings

Four organizations who were providing services to PLHIV were selected from Dhaka, Chittagong, Sylhet and Khulna of Bangladesh. These organizations helped researchers to get access of the participants as they do not disclose their HIV status in their community due to stigma.

### Study population

32 interviews were carried out with children (13-18years) and caregivers (10-13years) of CABA. Primary selection of participants was done through a list developed with the help of the organizations. 15 interviews were carried with children, whereas 17 were done with caregivers. For ethical considerations, caregivers of 10 to 13-year-old children were interviewed instead of the children. Participants were selected purposively based on their sex, age and location.

### Data Collection

Research associates including assistants with anthropology or social science background were recruited and trained to collect data. Training were provided on rapport building with the participants considering the sensitivity of the topic. Data was collected from March 2015 to May 2015.

In-depth Interviews (IDI) were conducted to understand the challenges faced by children and caregivers due to their HIV/AIDS status and HSPS role to support them. Desk review was conducted, which provided a framework for designing IDI guidelines.

The duration of interview was between 45 and 90 minutes. We stopped at 32 interview since we were not getting any new information from latest interviews. All the interviews were conducted at facility level settings to maintain the confidentiality with avoiding social backlash. Each interview was conducted in separate single room where no one could interrupt. Interviews were recorded through voice recorder. Field notes were also taken during data collection.

### Data Analysis

All the interviews were transcribed verbatim in Bangla by a qualitative research team. If there were any interruptions, field notes were used and merged to ensure data quality. After transcribing, data familiarization was done through transcript checking and reviewing.Thematic analysis was accomplished using a priori codes. They were then coded manually using a priori codes, sub-codes and emerging codes. Researchers involved with the study had frequent discussions to reflect and analyze data findings. Triangulation of data was done with the help of transcripts, field notes and researchers’ observations to ensure validity and reliability of the data.

### Ethical consideration

BRAC James P Grant School of Public Health research ethics committee approved the study. Written/verbal consents were taken from the participants. Study purpose, its benefits and risks, were explained to the participants. Informed consent was obtained from the parents and assent was obtained from the children. Permission was taken for notes taking and audio recording.Confidentiality and anonymity were maintained throughout the process. Since the participants were economically vulnerable, they were provided with money for lunch and compensation for transportation, which was a standard procedure for the organizations.

## Results

### Socio demographic profile of Caregivers (of 10-13 year old children)

Among the caregivers, 65% were male. Among all, 41% completed the primary level education. All the respondents were unemployed. Most respondents were receiving service for almost 5 years. A breakdown of background characteristics is presented in Tables 1.

**Table 1.** Socio-demographic profile of the caregivers interviewed

| **Variables** | **Frequency (percent)** |
| --- | --- |
| **Gender** |  |
| Male | 11 (64.7) |
| Female | 6 (35.3) |
| **District** |  |
| Dhaka | 3 (17.6) |
| Chittagong | 6 (35.3) |
| Khulna | 6 (35.3) |
| Sylhet | 2 (11.8) |
| **Education** |  |
| Primary school or less | 9 (52.9) |
| Secondary school | 1 (5.9) |
| Information not found | 7 (41.2) |
| **Work status** |  |
| Unemployed | 17 (100) |
| **Duration of receiving service** |  |
| More than 5 years | 9 (52.9) |
| Less than 5 years | 8 (47.1) |
| **HIV/AIDS status of the children** |  |
| Affected | 7 (41.2) |
| Infected | 10 (58.8) |

### Socio demographic profile of Children’s (13-<18 years of children)

Among child respondents, 67% were female. Almost half of the children were more than 15 years old. 47% children completed primary level education. A breakdown of background characteristics is presented in Tables 2.

**Table 2.** Socio demographic profile of the children interviewed

| **Variables** | **Frequency (percent)** |
| --- | --- |
| **Sex** |  |
| Male | 5 (33.3) |
| Female | 10 (66.7) |
| **Age** |  |
| 15-18 years | 7 (46.7) |
| 13-15 years | 8 (53.3) |
| **District** |  |
| Dhaka | 5 (33.3) |
| Chittagong | 2 (13.3) |
| Khulna | 6 (40) |
| Sylhet | 2 (13.3) |
| **Education** |  |
| Primary school or less | 4 (26.7) |
| Secondary school | 7 (46.7) |
| Information not found | 4 (26.7) |
| **Duration of receiving service** |  |
| More than 5 years | 3 (20) |
| Less than 5 years | 4 (26.7) |
| Don't know | 8 (53.3) |
| **HIV/AIDS status of child** |  |
| Affected | 11 (73.3) |
| Infected | 4 (26.7) |

## Challenges and vulnerability faced by children and caregivers

### Financial challenge

Beneficiaries highlighted poverty as a major challenge which prevented them from accessing services including the basic needs such as nutrition, education. CABA were vulnerable due to the risk of either having lost one/both parents or having parents who are ill and weak to engage in work. Both children and caregivers mentioned that the round-trip travel from their homes to facilities cost BDT 400-500 (USD $5-6) which they often found hard to manage.

> “*I took boat and then Rickshaw. After that I took Bus*…*it cost around 400tk* …*sometimes I take the money from father in law, sometimes from my elder sister in law*…*my brother gave me few times*” (Mother of a 10 year infected child)

Some participants mentioned that either they or their siblings had to quit education due to the death or illness of their parents.

> *“My brother has to quit his education as we have no one in our family who can income. After this, he joined a shop. Now, he earns money and sends to us. Every month he sends 4000-5000 tk. We have to struggle a lot*……*this money, you know, is not enough!”*
>
> (15 year HIV affected girl)

Beneficiaries also struggle to maintain their children nutrition due to poverty. Death of parents especially father increased financial vulnerability to children as in several occasions’ family inheritance were denied to them.

> *“People said that if the father died then his child does not get anything! Is that true? What will happen to my child? My father-in-law is old, what will happen when he dies? What if my brother-in-law does not give anything to him* [*my son*]*?”*(Mother of a 12 year old infected child)

### Gender norm

Gender issues played important role in vulnerability. Despite of availability of HSPS at NGOs serving PLHIV; women’s accessibility was restricted due to gender norms that prevent them from leaving home without permission. Women also had fewer employment opportunities, which leaves them dependent on male family members for money.

> *“As a woman I have encountered problems to travel here. They* [*family members*] *ask me about my every month visit. Sometimes they also remind me of travel expenses, it costs lots of money etc.”*(Mother of an 11-year-old HIV infected child)
>
> *“My husband died from this disease, after his death I live with my in laws. I am a woman; I am not allowed to leave home* [*for long distance travel*] *alone. Everyone ask me, where I am going? Why I am going? So, I bring my sister by telling lies to everyone.”* (Mother of a 12-year-old affected child)

Participants, especially girls, also mentioned being involving in household chores and taking care of the ill members of family.

> *“I am doing all the household chores since I was 6-year old. There was no one else in our house who could do it. My mother used to stay ill all the time. She could not do anything! I used to take care of her.”*(*14 year affected girl*)

Most caregivers also expressed worries about their children’s futures that reflect gendered expectations. For male children, the worries were mostly about their future employability, whereas for female children, caregivers were worried about their marriage ability. Low education level, and poor health condition also acted as challenges for women.

### Emotional wellbeing

Beneficiaries reported that HIV had negative effect on their lives. Significant sources of stress for caregivers and the children were dependency on others, financial instability, and the future impact of their disease on employability. Few mentioned that they were even feel suicidal. One caregiver expressed as,

> *When I feel bad, I cry. I feel that it’s better to die than bear life like this. I feel low, I hate myself*….*I am a burden on my parents with my kids. My parents married me off. My husband supposed to bear my cost, now his siblings supposed to support me* [*as respondent’s husband died from AIDS*], *I was supposed to have his assets to bear my expenses, but I have none & causing stress to my parents*. (HIV infected Mother of a 10 year infected child)

### Knowledge and social awareness on HIV/AIDS

Beneficiaries did not disclose their HIV status to anyone other than immediate family due to anxiety and constant fear of getting rejected and discriminated; which confirmed the persistent presence of HIV related stigma in communities and households.

> *They* [*Neighbours*] *said whoever would help her* [*my mother*] *would get the disease* [*AIDS*]…*They don’t visit us. She* [*Neighbour*] *told me, “I know, your mother has HIV. You should not share your utensils with her, nor should you share bed. This is contagious”*…*I started crying*. (15 year affected girl)
>
> “*I told the teachers about his illness status. Then one incident occurred. I went to a good school located in my area to admit my child, they did not admit him. They said, he could not do regular classes*…*They might help him to visit doctor, but they could not allow him to miss classes. Besides, this disease might spread from one child to ten other children. Then I told him, its ok, my child would remain in his current school.”*(Mother of 11 year infected child)

Some participants expressed their belief on God’s responsibility for the disease where man has no control over it.

> *“This is on God’s hand. Medication is a mere way*…*This is the fate of humans. Human’s wellbeing depends on Allah!!If he wants, people will get better with or without medicine. If he does not want, drug can’t do anything. Good or bad everything depends on him.”*(Mother of 12 year old infected child)

## Role of HSPS services in mitigating the challenges

### CCT and IGA

Beneficiaries identified CCTs and IGAs as much needed financial support they were receiving from the organizations and helped them to meet their basic needs. Children would not have continued their education had they not received this support. They also received allowance for buying books or school uniforms.

> *I stopped school when my mother died. My elder paternal uncle didn’t want to send me to school. If I would go to school it would hamper the household chores. However, I said, “I want to go to school.” I called my maternal uncle, he brought me here* [*the NGO providing HSPS*]. *They gave us money. Then, someone* (*name ommited0 from this NGO helped me to get admitted in a school. I received 24000tk* (*$300*) *from the NGO for my school admission. With that money I got admitted to a school, made my school uniforms and bought books. Someone* (*name omitted*) *from this NGO goes to our school to follow-up our progress*. (14 year affected girl)

Along with this, to promote the accessibility of the services organizations were helping the beneficiaries with the round-trip cost and lunch needed for the visits. In addition to these, nutrition support from these organizations also helped beneficiaries stay strong physically and mentally. However, these services got interrupted due to lack of funds. Beneficiaries identified this as one of the key services that had been discontinued because of lack of fund.

> *‘In the past I used to feel very weak and could not take care of my child*…*after receiving treatment and support from this organization I feel better. They used to give us money to buy nutritious food. They also gave us medicines and multivitamin for health problems like gastritis, skin infection, sometimes they used to give us biscuits, even milk* … *I feel much better now and can take care of my child’* (HIV infected mother of a 11 year old HIV infected child)

### Emotional support through counselling

Psychosocial support was one of the major services that were provided to the beneficiaries by the organizations. Counselling had positive impact on the emotional well-being of children and families. Beneficiaries received counselling in office or their homes, and during counselling, clients shared a range of social and emotional problems with providers. Counselling were given on information on HIV, life-style modification, how to take medications, nutrition, coping strategies of emotional/social stress etc. One HIV affected girl told that,

> *‘In the past I used to feel suicidal*…*why did this happen to my parents only* …*I haven’t heard anyone else get this disease apart from my parents*…*why did my father died*…*I would feel better if they* (*her parents*) *wouldn’t get infected by HIV*…*however, I have accepted this now*… *I talked with the sister* (*counsellor of the NGO providing HSPS service*)*”* (15-year-old HIV affected girl)

### Increased knowledge and social awareness

Community awareness programs were another key part of HSPS services; these programs are effective in the cultural context of Bangladesh as individual’s wellbeing in this culture is embedded in family’s wellbeing. Community sensitization through awareness raising programmes helped participants to gain acceptance in the community by reducing stigma.

> *“When my in-laws came to know about our illness, they immediately kicked us out of home*…*later people from here* [*the NGO providing HIV-sensitive protection services*] *spoke with them, they understood and we moved back in home”* (mother of a 12 year infected old child)

## Discussion

This article explored the challenges faced by CABA and how HSPS helped them to address those challenges. Several challenges were faced by CABA including financial, emotional and gender related. These challenges not only increased vulnerability of them but also affected the utilization of different services. Financial dependency and gender norm played significant role in vulnerability especially for women who often had weaker access to health services. HIV also hampered emotional wellbeing and caused enormous stress and depression among the CABA. Despite of several challenges HSPS helped CABA to cope up with the struggle through different services such as counselling, financial aid i.e. CCT and IGA etc. In addition to these, these services also helped to reduce stigma within families as well as communities through different awareness programmes.

Like our study it was found that lack of financial ability hindered basic needs and service accessibility of PLWHA (Dejman et al, 2015). CABA were at a higher risk of discontinuing their education and joining the work force. They faced unique financial pressures related to their parents’ illness or death. This often forced children to leave school and find paid work (Poulsen, 2006). Previous studies showed that financial support, even in small amount, helped to meet the basic needs such as food, shelter, school expenses and drugs along with promoting accessibility of services (UNICEF, 2007).

CCTs were a widely used intervention for improving the well-being of CABA. Global, regional, and local studies showed that CCT programs are an effective tool for improving child well-being which is similar to our study (Adato & Bassett, 2008; Baird et al., 2011; Nanda et al., 2014; Raynor & Wesson, 2006; Schady, 2006). An intervention done in Malawi among poor, HIV-affected households found that cash transfer of US$13/ month resulted in a 12% increase in overall school enrolment, a 5% increase in the number of students newly enrolled, and a 3% reduction in the dropout rate (Adato & Bassett, 2008).

At the same time, even highly cost-effective cash transfer programs were highly dependent on donor support, raising serious issues of sustainability. Donor dependency was a major issue for all four NGOs reviewed in this study, and affected the quality and sustainability of services. A review of cash transfer programs in Kenya, Malawi, and Zambia, found that the continuation of the vast majority of cash transfer programs were contingent on donor support and might not be appropriate in low income countries. (McCord, 2009).

Gender norms played crucial role in service accessibility especially for women as they often had weaker access to health services than men. Women’s autonomy is a significant predictor of a woman’s likelihood of seeking healthcare (Sen, 2007; Woldemicael & Tenkorang, 2010). Due to the subordinate role, it was found that women had to face worse economic disparities. They were more prone to the risk of quitting education, got evicted from home along with losing all financial support (Soskolne, 2003). In addition to these, girls became prone to early marriage as families often view early marriage as a way of protecting their daughters from harassment outside the home and ensuring their financial future (Field & Ambrus, 2008; Rashid, 2011).

Mental health disorder such as depression was common in HIV affected population & affected treatment adherence (WHO, 2017). Also, we found that rejection and discrimination related fear was consistent with HIV related stigma; which proved that psychosocial support for PLWHA was essential. Similar to our studies, studies conducted among HIV-infected adolescents in France found that peer supported adolescents suffered less anxiety about their illness and had a more positive perception of their treatment than adolescents who did not have peer support (Funck-Brentano et al., 2005). For both children and adult caregivers, social support from friends, family, and healthcare providers acted as a major determinant to seek care for their HIV-infected child. (Orner, 2006; Rouhani et al., 2017; Brouwer et al., 2000). The later study also found that much of the emotional stress experienced by caretakers were caused by economic instability, highlighting the importance of both psychosocial and economic support for caretakers.

UNICEF and other organizations emphasize the important of mobilizing and supporting community-based responses, especially efforts to reduce stigma and discrimination against PLWHA. Studies including ours suggested community sensitization can be a valuable tool reducing stigma and improving the lives of PLWHA. Sensitization training had a significant effect on attitudes and behaviour of the participants, also increased HIV knowledge particularly among community leaders. The evaluation found significant stigma reduction effects community wide (Nyblade, 2008; Apinundecha et al., 2007)

## Conclusion

This study demonstrates both the significant benefits of current service packages for CABA and substantial need for improvement and expansion of such services. Coordination and support from government is crucial as several challenges faced by CABA are structural, including insufficient incomes, inaccessibility of healthcare facilities, and limited educational opportunities. These obstacles prevent them from achieving their full potential and must be addressed through combined government and NGO actions. Also, expansion of existing economic assistance programs i.e. CCT, IGA are recommended as these are effective in improving child well-being. Employment and life skills training programs should be included in future to enable children to enter adulthood with the skills and knowledge necessary to thrive.

## Data Availability

The data that support the findings of this study are available from the corresponding author after publication of the manuscript in a peer review journal and on reasonable request.

## Abbreviations used

AIDS: Acquired immunodeficiency syndrome
ART: Anti-retroviral treatment
ARV: Anti-retroviral
CABA: Children affected or infected by HIV/AIDS
CCT: Conditional cash transfer
ERB: Ethics review board
HIV: Human Immunodeficiency Virus
HSPS: HIV-sensitive social protection services
IDI: In-depth Interviews
NGO: Non-governmental organization
PLHIV: Person living with HIV/AIDS
PLWHA: People living with HIV/AIDS
PMTCT: Prevention of mother to child transmission

## Acknowledgments

This study was completed with technical support from the UNICEF Bangladesh. Ashar Alo Society, Mukto Akash Bangladesh, Hope Care Center and Confidential Approach to Aids Prevention assisted us in data collection. National AIDS/STD Program (NASP) Bangladesh for provided overall guidance for this study. Dr. Sabina Faiz Rashid, Dean & Professor of BRAC JPGSPH, and Dr. Malabika Sarker, acting Dean & Professor of BRAC JPGSPH provided valuable comments on study methods.

## Author’s contributions

IM, TO & MZU conceptualized the research. IM was the PI. SC & TA supervised data collection. TA analysed the data under the guidance of IM & SC. TA wrote the first draft and improved it based on the feedback from IM. All authors read the final draft and approved it.

## Author’s information

TA worked as a deputy coordinator of MPH programme at BRAC James P Grant School of Public Health, BRAC University, Bangladesh. SC is PhD Candidate at Radboud University and Technical Advisor (Research) at BRAC James P Grant School of Public Health. TO was the former chief of HIV in UNICEF Bangladesh country office and currently is the UNICEF Deputy Representative in Pakistan. MZU is the HIV/AIDS specialist in UNICEF Bangladesh Country Office. IM is an Assistant Professor at Department of Public Health, College of Public Health and Health Informatics, Qassim University, Al Bukairiyah, Saudi Arabia and BRAC James P Grant School of Public Health, BRAC University, Bangladesh.

## Data Availability

The data that support the findings of this study are available from IM, the principal investigator of this study on reasonable request.

